# Sex and Gender Reporting and Female Participation Relative to Disease Burden in ClinicalTrials.gov

**DOI:** 10.64898/2026.11.14.26368113

**Authors:** Michael D. Green, Maryam Aziz, Rebecca D. Sullenger, Brandon J. Yoo, Emily C. O’Brien, Ann Marie Navar, Brenice Duroseau, Roland J. Thorpe, Carly L. Brantner

**Author notes:** Corresponding Author: Michael D. Green, Ph.D. Department of Health, Behavior and Society, Johns Hopkins Bloomberg School of Public Health. 615 N Wolfe St, Baltimore, MD 21205.

## Abstract

**Importance:** Assessing representation by sex assigned at birth and gender identity requires reporting of both in trials. If disclosure of full gender and sex participation from clinicaltrials.gov this is limited, it poses challenges in characterization and understanding of those participant experiences.

**Objective:** To determine how often ClinicalTrials.gov trials published trials report sex and gender, which characteristics are associated with reporting, and how participants’ female share compares with that of the general public.

**Design, Setting, and Participants:** Cross-sectional study of interventional trials registered to ClinicalTrials.gov, with or without US sites, with primary completion on or after September 27, 2008, and results first posted from January 1, 2009, through April 30, 2026.

**Exposures:** Funder type, phase, US sites, enrollment, primary completion year, number of countries, and therapeutic area.

**Main Outcomes and Measures:** Reporting of sex and gender, and the female participation-to-prevalence ratio (PPR; trial female share divided by the number of people with the condition).

**Results:** Among 58,007 trials (median enrollment, 72 [IQR, 31 to 206]; 42,644 [74%] with US sites), 553 (0.95%) reported gender and 57,683 (99.44%) reported sex. Gender reporting was more common with National Institutes of Health (NIH) funding (odds ratio [OR], 9.05; 95% CI, 6.65 to 12.6), US sites (OR, 4.21; 95% CI, 2.96 to 6.22), and larger enrollment. Across 10 conditions (15,552 mixed-sex trials), median PPR ranged from 0.68 (95% CI, 0.64 to 0.71) for chronic obstructive pulmonary disease to 1.05 (95% CI, 1.01 to 1.09) for obesity and had a relatively lower female share in 8. Among the 10 conditions most prevalent in our cohort, 6,917 of 16,097 trial condition combinations (43.0%) reached a PPR of at least 0.90; odds of reaching it were higher for NIH-funded (OR, 1.22; 95% CI, 1.11 to 1.34) and lower for government-funded trials (OR, 0.26; 95% CI, 0.20 to 0.33) as compared to industry-funded trials.

**Conclusions and Relevance:** In ClinicalTrials.gov, sex reporting is nearly universal, whereas gender reporting is concentrated among NIH-funded, US-based, and larger trials. Female trial enrollment fell below the female share of people with the condition for most conditions, emphasizing the benefit of condition-specific benchmarks in clinical trials.

## INTRODUCTION

Representative enrollment of sociodemographic groups in clinical trials that reflects the people affected by the disease supports the generalizability of results and increases public trust in research.^1^ Despite this, clinical trial enrollment does not always reflect target populations by sex assigned at birth or gender identity. Women have historically been underrepresented in clinical research.^2,3^ Sexual and gender minority populations, a group who experience many documented health disparities, also appear underrepresented in clinical trials.^4^ Recently, United States federal health agencies like the National Institutes of Health (NIH) have implemented the Sex as a Biological Variable policy (NOT-OD-15-102),^5^ emphasizing the purpose of having biomedical research across multiple sexes to increase generalizability of clinical research. This action builds upon earlier initiatives, like the 1993 NIH Revitalization Act,^6^ which sought to reverse the practice of excluding women from biomedical research. After this multi-decade push to include women in research (and increase the generalizability of biomedical studies), the public would benefit from an update on any progress on the goals of these initiatives.

Representation of women in clinical research varies by study type and disease.^7^ Women are more often underrepresented in interventional than in observational studies, partly because of exclusion criteria such as pregnancy.^8^ Studies comparing the female enrollment of trial participants with those with the disease found women underrepresented in some oncology^9,10^ and cardiovascular trials.^11–13^ As a result, clinical decisions for women may rest on observational data instead of trial evidence.^14^ ClinicalTrials.gov is the largest registry of clinical trials in the United States, and is the central location where sponsors submit trial details and report results.

Assessing representativeness requires accurate, distinct reporting of sex and gender, because each may influence disease risk, clinical presentation, and response to treatment.^15^ Most trials report participant sex in their results,^16^ but gender reporting is less common. The terms are sometimes used interchangeably: the ClinicalTrials.gov gender eligibility description field is intended to capture self-identified gender,^17–19^ but its entries may reflect participants’ sex or other information rather than gender identity.

To address these gaps, we examined trials with results posted on ClinicalTrials.gov to describe sex and gender reporting and its correlates, compare female enrollment in common conditions with each condition’s female share, and identify trial characteristics associated with reaching at least 90% of that share.

## METHODS

### Trial dataset and data extraction

We queried the ClinicalTrials.gov application programming interface for interventional trials with posted results (summary results publicly posted on ClinicalTrials.gov) through April 30, 2026, excluded terminated trials to avoid incomplete or inconsistently reported demographics, and linked them to Aggregate Analysis of ClinicalTrials.gov (AACT) flat files on the NCT identifier. Funder type (industry, National Institutes of Health [NIH], government [US federal and other public], or other) came from the lead sponsor and collaborators. Our data retrieval and linkage approach are described in eMethods 1.

For 575 trials with a customized or multigroup baseline sex or gender table, which can have nested tables or have an inconsistently structured count of participants across trial arms,^20^ a large language model (Claude Haiku 4.5, accessed May 2026) parsed the information and one reviewer checked every parse against the online ClinicalTrials.gov registry record: 475 (82.6%) were correct and 100 had a format or count problem. The reviewer (B.Y.) supplied corrected values for 98, which the analyses use; the other 2 keep their extracted values, flagged (eMethods 2). We report the LLM-assisted extraction and the AI-assisted classification following the TRIPOD-LLM reporting guideline (eAppendix 1).^21^ Our study was deemed exempt by the Duke University Health System Institutional Review Board (IRB #Pro00120645).

### Analytic cohorts

The primary analytic cohort was comprised of trials with a primary completion date on or after September 27, 2008. This is to align with when submission of summary results became required for applicable clinical trials under Section 801 of the Food and Drug Administration Amendments Act (FDAAA 801).^22^ We restricted the cohort to trials whose results were first posted on or after January 1, 2009, the start of the study window (1 trial excluded; eTable 2). We excluded 755 AACT records absent from our application programming interface query whose results were posted after April 30, 2026 (eTable 2). We also analyzed a secondary mixed-sex analytic cohort comprising trials with eligibility open to multiple sexes, not based on gender identity, which reported the number of female and male participants enrolled.

### Comparison populations

For each condition, the comparison population were people in the United States with that condition, and their female share was the benchmark (Outcomes). We studied the 10 conditions with the most mixed-sex trials, treating each MeSH condition heading as its own therapeutic area except all neoplasm headings formed 1 condition and all diabetes mellitus headings another. A trial contributed 1 trial condition combination for each of these conditions it carried. Because the diabetes and neoplasm benchmarks describe type 2 diabetes and malignant cancer, a trial did not count toward diabetes when its headings named another diabetes type, or only prediabetes, and no type 2 heading (543 of 2,634), or toward neoplasms when MeSH filed its neoplasm headings only under cysts, hamartomas, or precancerous conditions (145 of 8,892) (eMethods 4; Supplement 2, eTable 1).

### Outcomes

The 3 primary outcomes were reporting of sex, reporting of gender, and female participation relative to each condition’s benchmark. Following published definitions and the Sex and Gender Equity in Research (SAGER) guidelines,^23^ sex refers to biological attributes (chromosomes, hormones, and anatomy), usually recorded as sex assigned at birth and not always binary; and gender refers to socially constructed roles, behaviors, and identities, including gender identity.^24^The ClinicalTrials.gov registry tables do not show how either was ascertained. We defined a trial to have reported sex if it posted female and male counts under any measure label. We defined a trial as reporting gender if it posted gender-diverse categories, or cisgender and transgender categories (gender-titled measures with only female and male [or woman and man] counts were classified as sex, so gender reporting as defined in this study provides a lower bound on collection of gender identity). We classified every measure into 1 of 3 states: a breakdown was reported, participants were recorded only as unknown or not reported, or the measure was absent. A breakdown covering only some participants, or only clusters, was flagged as incomplete composition (eMethods 2).

When sex was reported and within the mixed-sex cohort, we defined percent female as female participants divided by female plus male participants, so the denominator excluded participants of unknown sex. To evaluate representativeness of women in specific conditions, we used the female participation-to-prevalence ratio (PPR), which divides a trial’s female share by the female share of people with the condition (the benchmark); a PPR of 1 means female trial enrollment equaled the benchmark. We calculated a PPR for each trial condition combination and summarized each condition by the median PPR, which equals the median trial female share divided by the benchmark. For 8 conditions, the benchmark was the female share of diagnosed patients in PCORnet,^25^ a care-seeking population, from Population Insights reports or a 2023 network-wide query. Benchmarks for chronic obstructive pulmonary disease (COPD) and pain were defined as the female share of adults reporting COPD and chronic pain, respectively, in the 2023 National Health Interview Survey (NHIS)^26–27^ (eMethods 3; eTable 3). We assigned trials to conditions based on each trial’s MeSH headings resolved to MeSH 2026 descriptors.^16,28,29^

### Therapeutic-area classification and clinical review

The 10 therapeutic-area indicators are the 10 most common MeSH disease branches among the 58,007 cohort trials (Supplement 2, eTable 1). For our analysis a trial is in a therapeutic area when any heading falls under the branch’s top-level tree [e.g. Neoplasms (C04)], with one exception [the descriptor Depression (D003863), which MeSH files under Behavior and Behavior Mechanisms (F01) rather than Mental Disorders (F03)]; through clinical judgment, we also placed it in mental health. The therapeutic areas are broad (153 to 622 distinct descriptors each) and overlap. Many MeSH descriptors have more than one parent category. For example multiple myeloma places 593 trials in both oncology and cardiovascular, and non-small-cell lung carcinoma 903 in both oncology and respiratory (Supplement 2, eTable 1).

To check our assignments, 2 clinicians (B.D. and R.D.S.) independently reviewed 100 randomly sampled mappings: 56 therapeutic-area assignments across the 10 areas, 40 condition assignments, and 4 decisions under the diabetes and neoplasm exclusions. For each mapping, they saw the trial record and our assignment and answered agree, disagree, or cannot tell; a third clinician (A.M.N.) adjudicated every mapping without 2 agree or 2 disagree answers, including mappings either clinician answered cannot tell. The review reports how often clinicians confirmed the displayed assignments (Supplement 2, eFigure 1, Part C).

**Figure 1.**
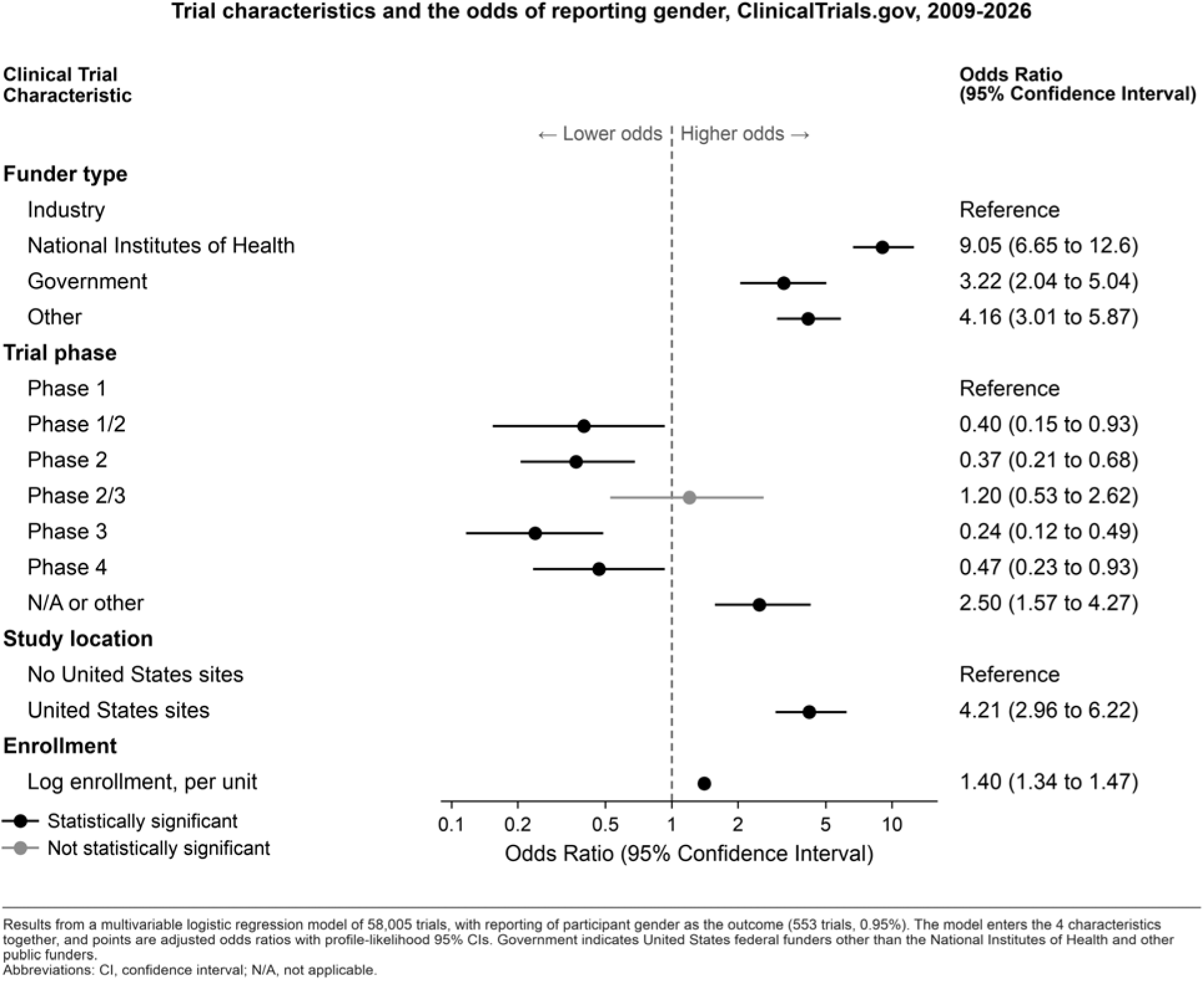
Trial characteristics and the odds of reporting gender, ClinicalTrials.gov, 2009-2026 Results from a multivariable logistic regression model of 58,005 trials, with reporting of participant gender as the outcome (553 trials, 0.95%). Created with Claude Science (beta; Anthropic), using Claude Opus 5.5, and Matplotlib 3.11.1. Abbreviations: CI, confidence interval; N/A, not applicable.

### Statistical analysis

For reporting gender, we fit a multivariable logistic regression on funder type, trial phase, US study sites, and log enrollment in complete cases. Median PPRs carry bootstrap percentile 95% CIs from 2,000 resamples of each condition’s trials. For reaching a PPR of at least 0.90, we fit a logistic regression of trial-condition combinations, with predictors of funder type, phase, US sites, primary completion year, log enrollment, number of countries, and 10 nonexclusive therapeutic-area indicators, and with standard errors clustered by trial and Wald 95% CIs. Analyses used R (R Foundation for Statistical Computing)^30^ and Python^31^ with the sandwich package (version 3.1-3)^32^ for clustered standard errors and the boot package (version 1.3-28)^33^ for bootstrap intervals (eMethods 1); Wald P values are 2-sided.

## RESULTS

### Reporting of sex and gender

Of the 58,007 trials in the primary cohort (Table 1), only 553 (0.95%) reported participant gender, whereas 57,683 (99.44%) reported sex. The primary cohort included 30,922 industry-funded trials (53%) and 9,650 NIH-funded trials (17%); 42,644 (74%) had at least one US site, and median enrollment was 72 participants (IQR, 31 to 206). As mutually exclusive states (Table 1; by funder type, eTable 1), 57,168 trials (98.6%) reported sex only, 515 (0.89%) reported both sex and gender, 38 (0.07%) reported gender only, and 286 (0.49%) reported neither. Of those reporting neither, 173 recorded participants only as unknown or not reported or stated sex was not collected, 71 had no sex or gender measure, and 42 posted a measure without a usable breakdown.

**Table 1.** Baseline Characteristics of the Primary Clinical Trial Cohort.

| Characteristic | N = 58,007 |
|---|---|
| Funder type, No. (%) |  |
| Industry | 30,922 (53) |
| NIH | 9,650 (17) |
| Government (US federal and other public) | 2,532 (4.4) |
| Other | 14,903 (26) |
| Trial phase, No. (%) |  |
| Phase 1 | 4,862 (8.4) |
| Phase 1/2 | 2,803 (4.8) |
| Phase 2 | 13,818 (24) |
| Phase 2/3 | 1,008 (1.7) |
| Phase 3 | 10,340 (18) |
| Phase 4 | 6,108 (11) |
| Not applicable or other | 19,068 (33) |
| Study location, No. (%) |  |
| No US sites | 15,363 (26) |
| US sites | 42,644 (74) |
| Enrollment, median (IQR), participants | 72 (31 to 206) |
| Unknown | 2 |
| Time to report results, median (IQR), months<sup>a</sup> | 20 (14 to 37) |
| Sex and gender reporting, No. (%)<sup>b</sup> |  |
| Sex only | 57,168 (99) |
| Both sex and gender | 515 (0.89) |
| Gender only | 38 (0.07) |
| Neither sex nor gender | 286 (0.49) |
| Recorded only as unknown or not reported, or not collected | 173 (0.30) |
| No sex or gender measure | 71 (0.12) |
| Measure without a usable female and male breakdown | 42 (0.07) |
| Condition category (one per trial), No. (%)<sup>c</sup> |  |
| Neoplasms | 11,212 (19) |
| Diabetes mellitus | 2,733 (4.7) |
| Depression | 880 (1.5) |
| Pain | 795 (1.4) |
| Obesity | 741 (1.3) |
| Asthma | 634 (1.1) |
| Chronic obstructive pulmonary disease | 588 (1.0) |
| HIV | 575 (1.0) |
| COVID-19 | 567 (1.0) |
| Schizophrenia | 537 (0.9) |
| Other condition (outside the 10 above) | 29,711 (51) |
| No MeSH condition heading | 9,034 (16) |

<sup>a</sup> From primary completion to first results posting; 223 trials had no computable interval (negative after a completion-date revision, or a missing date).
<sup>b</sup> Mutually exclusive reporting states (definitions in Methods); the 3 rows under Neither are reported separately.
<sup>c</sup> Each trial's single most prevalent MeSH condition category; categories are mutually exclusive and differ from the Figure 2 rows.
Abbreviations: IQR, interquartile range; MeSH, Medical Subject Headings; NIH, National Institutes of Health.

Among the 58,005 trials with complete covariates, gender reporting was more common with NIH than industry funding (OR, 9.05; 95% CI, 6.65 to 12.6), with US sites (OR, 4.21; 95% CI, 2.96 to 6.22), and with larger enrollment (OR per log unit, 1.40; 95% CI, 1.34 to 1.47) (Figure 1). Of the 553 gender-reporting trials, 524 counted at least 1 participant in a gender-minority or self-described category, most often nonbinary or gender-expansive (4,711 participants in 222 trials) or transgender (4,473 participants in 199 trials). Counting only categories which clearly denote a gender minority and so excluding “another or self-described” (which includes categories labeled only “Other”), 360 trials reported at least 9,492 gender-minority participants. Intersex (less than 50 participants in 12 trials), cisgender-labeled (19,114 participants in 38 trials), and explicit unknown, not reported, or declined categories (11,673 participants in 154 trials) were counted separately, and never as gender diverse (eTable 4).

Gender reporting rose from 0.27% of trials completing in 2017 to 3.47% of those completing in 2024 (1.75% pooled from 2017, approximating FDAAA Final Rule^22^ coverage). Of trials completing in 2025, 25 of 783 (3.19%) reported gender, and none of the 19 completing in 2026 did. Recent completion years are incomplete based on how long trials completed in 2016 through 2020 took to post results (eMethods 1). When trials were instead grouped by the year results were first posted, gender reporting was 2.77% in 2024, 3.02% in 2025, and 2.57% in January through April 2026.

### Female participation overall and by condition

Among the 50,401 mixed-sex trials, mean percent female was 48.2%, ranging by primary completion year (2009 to 2025) from 46.4% (2012) to 51.0% (2024) (Figure 4). Female-only trials outnumbered male-only trials in every completion year, 4,436 vs 2,208 in total (Figure 4).

The 10 most common conditions we analyzed covered 15,552 trials and 16,097 trial-condition combinations (Figure 2; eTable 5). Median PPR was below 1, with the 95% CI below 1, in 8 of the 10 conditions, meaning the median trial enrolled a smaller female share than the benchmark; values ranged from 0.68 (95% CI, 0.64 to 0.71) for COPD to 0.94 (95% CI, 0.92 to 0.97) for asthma. In obesity trials, the median PPR was above 1 (1.05; 95% CI, 1.01 to 1.09), meaning the median trial’s female share exceeded the female share of PCORnet patients with obesity (61.1%). For pain, the 10th condition (748 trials), the median PPR was 1.01 (95% CI, 0.99 to 1.04) against a chronic-pain benchmark.

**Figure 2.**
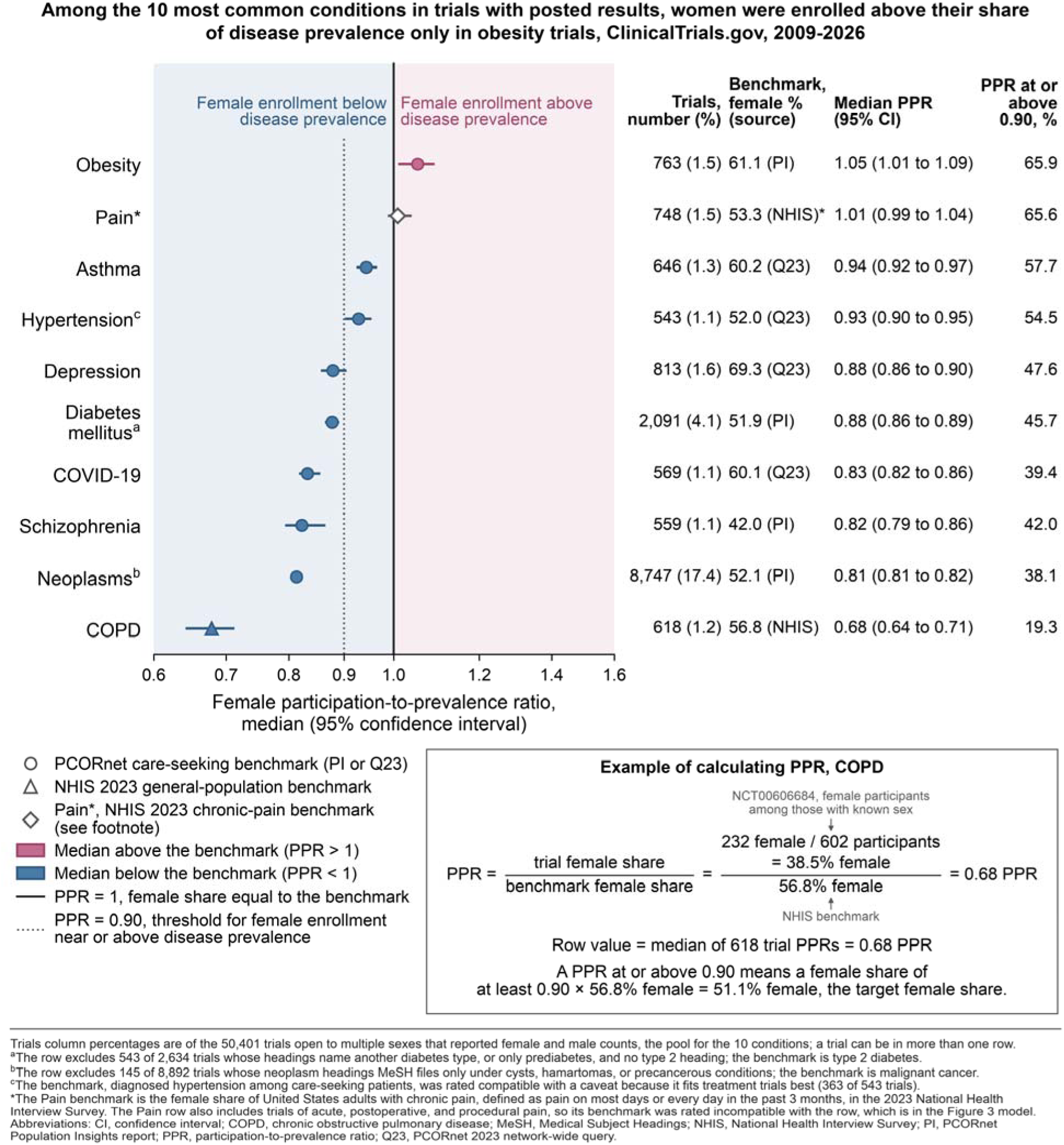
Among the 10 most common conditions in trials with posted results, women were enrolled above their share of disease prevalence only in obesity trials, ClinicalTrials.gov, 2009-2026. Rows are the 10 condition categories with the most trials open to multiple sexes, counting a trial under every category it carries and keeping one neoplasms row. The last column gives the percentage of trials whose female share reached at least 90% of the benchmark (PPR ≥ 0.90). Created with Claude Science (beta; Anthropic), using Claude Opus 5.5, and Matplotlib 3.11.1. Abbreviations: CI, confidence interval; COPD, chronic obstructive pulmonary disease; NHIS, National Health Interview Survey; PI, PCORnet Population Insights report; PPR, participation-to-prevalence ratio; Q23, PCORnet 2023 network-wide query.

### Trial characteristics associated with reaching the PPR threshold

Overall, 6,917 of 16,097 trial-condition combinations (43.0%), from 15,552 trials, reached a PPR of at least 0.90, meaning a female share at least 90% of the benchmark. Adjusted odds of reaching the PPR of 0.90 (Figure 3) were higher for NIH-funded (OR, 1.22; 95% CI, 1.11 to 1.34) and other-sponsor trials (OR, 1.16; 95% CI, 1.05 to 1.27) than for industry trials, lower for government-funded trials (OR, 0.26; 95% CI, 0.20 to 0.33), and higher for trials with US sites (OR, 1.38; 95% CI, 1.26 to 1.51), without a labeled phase, and with later completion dates.

**Figure 3.**
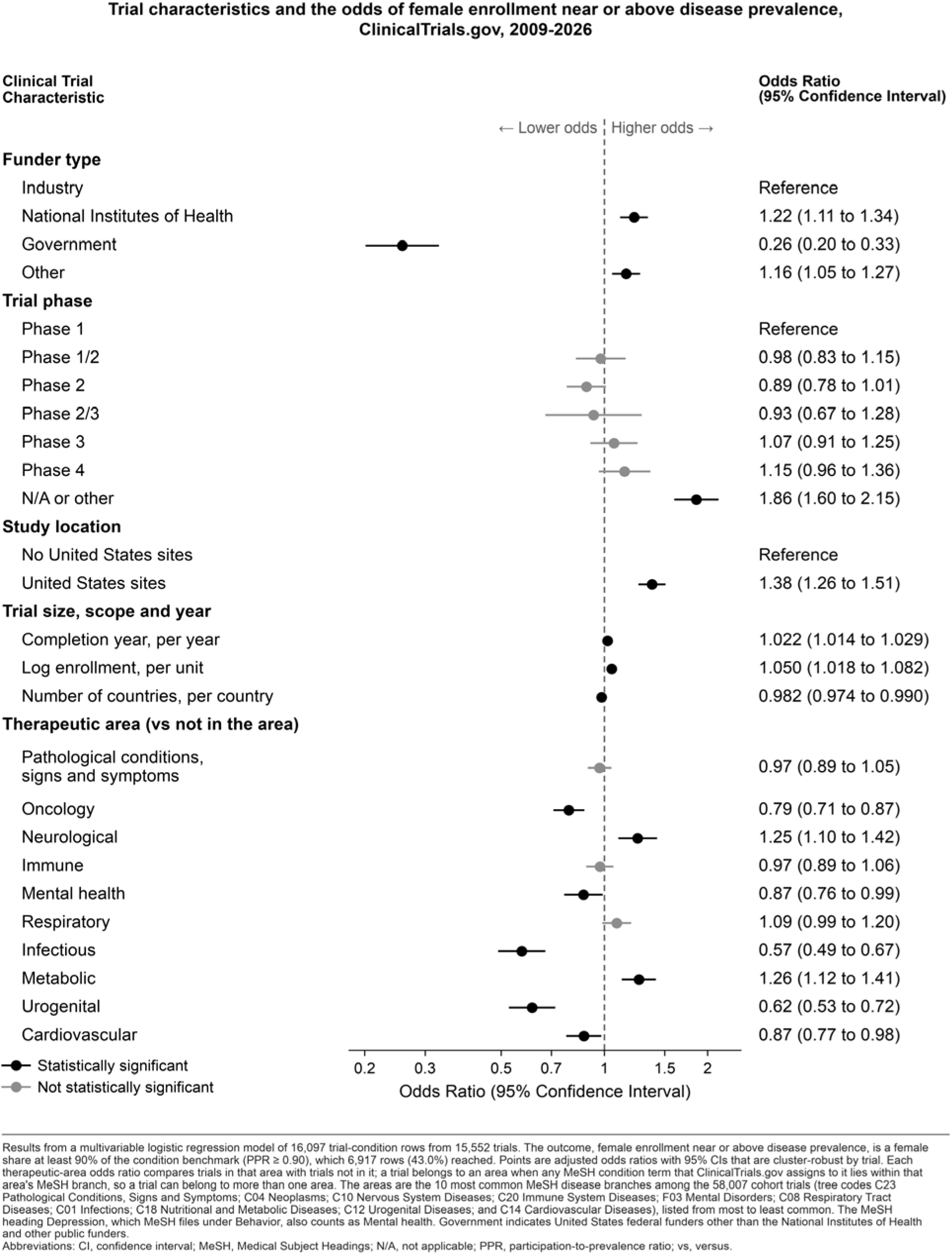
Trial characteristics and the odds of female enrollment near or above disease prevalence, ClinicalTrials.gov, 2009-2026. Results from a multivariable logistic regression model of 16,097 trial-condition combinations from 15,552 trials. Outcome is female enrollment near or above disease prevalence, is a female share at least 90% of the condition benchmark (PPR ≥ 0.90), which 6,917 (43.0%) reached. Each therapeutic-area odds ratio compares trials in that area with trials not in it. A trial in several conditions contributes a combination for each (487 trials), so the model clusters standard errors by trial and reports Wald 95% CIs. Created with Claude Science (beta; Anthropic), using Claude Opus 5.5, and Matplotlib 3.11.1. Abbreviations: CI, confidence interval; MeSH, Medical Subject Headings; N/A, not applicable; PPR, participation-to-prevalence ratio; vs, versus.

**Figure 4.**
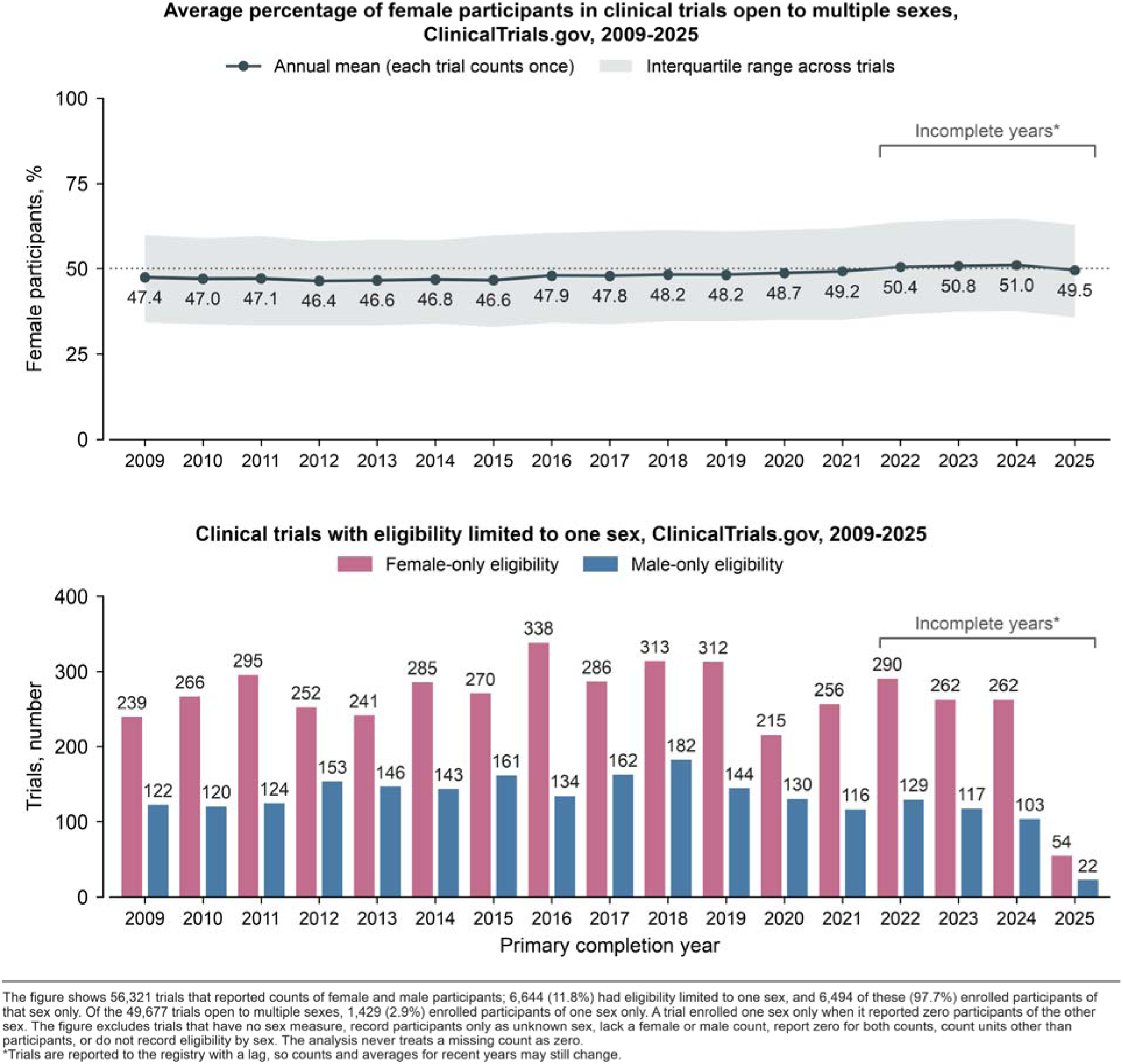
Female representation and single-sex trials by primary completion year, ClinicalTrials.gov, 2009-2025. The top panel shows the trials open to multiple sexes, with the annual mean of within-trial percent female among participants with known sex, each trial counted once, and the interquartile range across trials. The bottom panel shows the trials with female-only eligibility (4,436) or male-only eligibility (2,208. 56,321 trials reported counts of female and male participants; 6,644 (11.8%) had eligibility limited to one sex, and 6,494 of these (97.7%) enrolled participants of that sex only. Of the 49,677 trials open to multiple sexes, 1,429 (2.9%) enrolled participants of one sex only. Created with Claude Science (beta; Anthropic), using Claude Opus 5.5, and Matplotlib 3.11.1.

## DISCUSSION

Our study examined representativeness by sex and gender by whether they report the characteristic and who is enrolled among the studies that reported these characteristics. Gender is largely absent from ClinicalTrials.gov results, so the registry cannot yet show whether individuals with minority gender identities are included widely in clinical trials in the US. The 2016 FDAAA Final Rule requires a sex/gender baseline measure which can satisfied with a male female account^22^ and the register therefore shows little on gender beyond binary sex. NIH-funded trials (subject to NIH policies on the inclusion of women and on sex as a biological variable)^5^ were the most likely to report gender. Registry data alone cannot show whether those policies explain the difference in reporting. When evaluating this data cumulatively, we found male/female participation in clinical research is near even participation percentages, but our more granular analysis examining common diseases and therapeutic areas provides further detail. An aggregate female share close to 50% concealed shortfalls in the participation of women relative to their share of people with each condition. The median PPR was below 0.90 in 6 of the 10 conditions we analyzed, and only obesity trials enrolled a female share above the benchmark with a 95% CI above 1.

While we are optimistic about the progress in reporting of presumed biological sex, that alone does not fully capture representativeness with respect to sex and gender. The conditions furthest below their benchmarks show where our medical research ecosystem could allocate more resources and efforts towards increasing the share of other gender groups. Improving the representation of women and gender-diverse people requires trials designed around participants’ lives: critically reviewed eligibility criteria, flexible or decentralized visits, reimbursement of transportation, caregiving, and other costs, and culturally and linguistically responsive recruitment.^34–37^ The SAGER guidelines recommend researchers distinguish sex and gender, incorporate them into design and analysis when relevant, and justify exclusions explicitly.^23,24^ Investigators can also adopt a presumption of eligibility for people who are pregnant, lactating, or capable of pregnancy, requiring scientific justification for exclusion.^38^ For transgender and gender-diverse people, inclusion extends beyond a demographic checkbox to community co-design of research questions, measures, and recruitment, and registry reporting that separates sex assigned at birth from gender identity would keep these participants scientifically visible.^39,40^

### Limitations

Our analysis should be considered alongside some limitations. First, trials that never post results are not represented, and because posting lags behind completion, recent completion years reflect faster-reporting trials. Second, we classified each baseline table as submitted, so trials splitting sex and gender across tables may be undercounted as reporting both. Third, ClinicalTrials.gov predominantly registers trials with US involvement, so this study represents a US-centric view, and registries which examine global trials and their equivalent reporting standards might find different results around the inclusion of women and gender.

Finally, we note that our calculated PPRs depend on benchmarks which mix care-seeking (PCORnet) and general-population (NHIS) sources, which are all from the United States. PCORnet denominators include children, so the adult-eligible analysis (eTable 4) restricts the trials but not the benchmark. Our clinical categories are approximations: a National Library of Medicine algorithm assigns MeSH headings from free-text condition fields, and that algorithm differs from clinical review and has not been validated for this purpose. The pain benchmark describes US adults with chronic pain, whereas the broad Pain heading also covers acute, postoperative, and procedural pain trials, so the pain PPR, and the Pain row’s contribution to the Figure 3 model, may misstate female participation in pain trials. Furthermore, 8 benchmarks describe diagnosed patients in care at PCORnet sites, who lean female (56.4% of the 2023 any-diagnosis cohort). Our care-seeking benchmark can therefore exceed the female share of everyone with the condition, so we emphasize that the PPRs should be interpreted with this caveat in mind. The neoplasm benchmark is a treated prevalence which includes long-term survivors, so sex differences in survival may affect it, and prevention trials enroll people without the condition; our exclusion rules narrow these mismatches, but discrepancies like unnamed diabetes types and heuristically classed neoplasm headings remain.

### Conclusions

Trials with posted results on ClinicalTrials.gov almost always report how many females and males enrolled but rarely report participant gender. Female enrollment in mixed-sex trials were close to 50% overall but fell below the female share of people with the condition in most conditions with a compatible estimated benchmark. Separating sex assigned at birth from gender identity in registry reporting and comparing enrollment with each condition’s female share rather than an even split, facilitates sponsors, regulators, and clinicians in evaluating whether trial evidence applies to the patients they ultimately treat.

## Supporting information

Supplement 2. MeSH Heading Classification of Trials

Supplement 1. Supplementary Online Content

## ARTICLE INFORMATION

### Author Contributions

Michael D. Green and Carly L. Brantner had full access to all the data in the study and take responsibility for the integrity of the data and the accuracy of the data analysis. The CRediT author statement is as follows: **Michael D. Green:** Conceptualization, Methodology, Software, Formal analysis, Investigation, Resources, Data Curation, Writing - Original Draft, Writing - Review & Editing, Funding acquisition. **Maryam Aziz:** Data Curation, Writing - Original Draft, Writing - Review & Editing. **Rebecca D. Sullenger:** Data Curation, Writing - Original Draft, Writing - Review & Editing. **Brandon J. Yoo:** Data Curation, Writing - Review & Editing. **Emily C. O’Brien:** Conceptualization, Methodology, Writing - Original Draft, Writing - Review & Editing, Supervision. **Ann Marie Navar:** Conceptualization, Methodology, Writing - Original Draft, Writing - Review & Editing, Supervision. **Brenice Duroseau:** Data Curation, Writing - Original Draft, Writing - Review & Editing. Roland **J. Thorpe Jr:** Writing - Review & Editing, Supervision. **Carly L. Brantner:** Conceptualization, Methodology, Validation, Formal analysis, Writing - Original Draft, Writing - Review & Editing.

### Data Sharing Statement

Aggregate Analysis of ClinicalTrials.gov (AACT) data can be downloaded at https://aact.ctti-clinicaltrials.org/. Data for condition prevalence benchmarks were in part pulled from a 2023 PCORnet^®^ query, as well as publicly available Population Insights reports. PCORnet has been developed with funding from the Patient-Centered Outcomes Research Institute^®^ (PCORI^®^). The statements presented in this work are solely the responsibility of the author(s) and do not necessarily represent the views of organizations participating in, collaborating with, or funding PCORnet or of PCORI.

### Use of AI

One author (M.G.) used a large language model (Claude Opus 4.8, 5.0 and Fable 5.0; Anthropic) in June-August 2026 to assist with code review, debugging, and validation of the initial analysis. Two authors reviewed and verified all code and outputs during the first draft stage (M.G. and C.B.). In the initial draft Claude Opus 4.8 and 5.0 were used by M.G. to outline headings and correct the grammar of sentences as well as present potential critiques of the argument presented for the article (M.G.). In September and October 2026, Fable 5.1, Opus 5.0, and Opus 5.5 were used within the Claude Science beta harness to edit the analysis pipeline and build the MeSH classification audit file. In October and November 2026, the same harness, running Claude Opus 5.5, generated Figures 1 to 4 and the diagrams in eFigure 1, Parts A to C (Supplement 2), with the Python library Matplotlib (version 3.11.1), under M.G.’s direction. In October 2026 OpenAI’s Codex Astra 6 was used to design a web app (M.G.) for review of sorting of trials into therapeutic areas, in every case under M.G.’s direction and with each change reviewed by M.G. before acceptance. Finally, there was a step described within the methods where an LLM (Claude Haiku 4.5; Anthropic; accessed May 2026) was used by M.A. to take a first pass at parsing information on customized trial categories, then a human verified them (B.Y. under supervision of M.G.). In October 2026, under M.G.’s direction, Claude Sonnet 5, Sonnet 5.5 and Opus 5.5 answered the 100 clinician review questions (October 25), and Claude Sonnet 5.5 and Opus 5.5 checked a sample of the MeSH-based assignments (October 30 and 31); no estimate uses these answers (Supplement 2). Supplement 1, eAppendix 1 gives the TRIPOD-LLM checklist. All authors take responsibility for the integrity of AI-assisted content.

### Additional Contributions

N/A

