## Supplement 2. MeSH Heading Classification of Trials for "Sex and Gender Reporting and Female Participation Relative to Disease Burden in ClinicalTrials.gov"

eMethods. Checks of the Therapeutic-Area and Condition-Group Assignments

eFigure 1. Therapeutic-Area Classification: Part A, MeSH Rule; Part B, AI Check of the Part A Assignments; Part C, Clinical Review

eTable 1. Therapeutic Areas and Condition Categories as Parent MeSH Categories: Breadth and AI Check of Assignments

eTable 2. Clinician Review: Independent Answers and Final Outcomes

eTable 3. Two AI Review Chains on 1,000 Further Assignments: Final Decisions and Auditor Revisions

This supplementary material has been provided by the authors to give readers additional information about their work.

**eMethods. Checks of the Therapeutic-Area and Condition-Group Assignments**

The MeSH rules in eFigure 1, Part A assign each trial to therapeutic areas and condition groups. We checked these assignments in 2 ways. A review of 100 assignments by clinicians and checks of other sampled assignments by large language models.

**1. Clinician review**

Two clinicians (B.D. and R.D.S.) independently judged 100 assignments, each in a different trial with posted results, sampled at random within 24 strata: 56 therapeutic-area assignments (5 or 6 per area), 40 condition-group assignments (4 per condition), and 4 decisions under the diabetes-type and nonmalignant-neoplasm rules. Each clinician read the reviewer guide and 3 worked examples, then saw each trial’s registry information and the displayed assignment, and answered Agree, Disagree (with a reason), or Cannot tell. Two Agree or Two Disagree answers settled a question. Differing answers went to a third clinician (A.M.N.), who saw both clinicians’ answers and notes, and recorded Agree or Disagree or left the question unresolved.

After recording their decisions, the adjudicator asked us to check their comments because they were unsure what Agree and Disagree referred to. We restated the definitions, including that a trial can belong to more than 1 category, and asked them to keep or change the 7 Disagree decisions whose comments proposed a different category, without suggesting an answer. They changed 5 to Agree. We report the results with these 5 changes and as first recorded. For each family we report the numbers confirmed, not confirmed, and unresolved, the 2 clinicians’ exact agreement, and unweighted Cohen κ (eTable 2).

**Results.** Both clinicians gave the same answer to 46 of 56 therapeutic-area questions (82.1%; κ = 0.67) and 35 of 40 condition-group questions (87.5%; κ = 0.46); 17 questions went to the adjudicator. Clinicians confirmed 31 of 56 therapeutic-area assignments (55.4%; as first recorded, 28, 50.0%), from 0 of 5 for Urogenital to 6 of 6 for Oncology; 36 of 40 condition-group assignments (90.0%; as first recorded, 34, 85.0%), with 3 not confirmed and 1 unresolved, the question both clinicians answered Cannot tell; and all 4 inclusion or exclusion decisions. Of the 25 therapeutic-area assignments not confirmed, 23 were questions both clinicians answered Disagree; across their 46 answers the reasons were “not the condition studied” (23), “cancer origin, spread or course only” (14), “MeSH placement is misleading” (4), and another reason (5).

**2. Review by large language models**

On 30 and 31 October 2026, 2 AI models checked every therapeutic-area and condition-group assignment of 1,091 trials, drawn at random within 19 strata from the 15,454 Figure 3 trials outside the clinician sample. claude-sonnet-5-5 (Anthropic) was provided each trial’s titles, summaries, registered conditions, interventions and MeSH headings, with each assignment and the rule that made it. It then answered supported, not supported (quoting the registry record), or insufficient evidence for each assignment. Without seeing those answers, claude-opus-5-5 repeated the check for 378 of the 388 trials in which the first model called at least 1 assignment not supported or insufficient evidence, and for 100 control trials in which it had supported every assignment. Each step allowed 1 retry per trial; missing outputs were never filled.

**Results.** 1,070 of 1,091 first-pass checks were valid; 21 remained missing. The first model marked 520 of 2,997 answered assignments (17.4%) not supported (495 therapeutic-area and 25 condition-group assignments); the second model answered 505 of these and also marked 449 not supported, and it supported all 216 assignments in the 100 control trials. By therapeutic area, the share marked not supported ranged from 2.6% (Oncology) to 75.0% (Urogenital); no condition group exceeded 4.1% not supported (eTable 1, panel B; eFigure 1, Part B).

On 3 and 4 November 2026, 2 AI review chains judged 1,000 further assignments from the Figure 3 frame: 50 drawn at random in each of the 10 therapeutic areas and 10 condition groups, after excluding the 1,191 trials used in the clinician review and in the check above, so the sample shares no trial with either. Each unit showed 1 assignment of a trial and the rule behind it. In chain A, claude-haiku-5-5 judged the assignment and claude-sonnet-5-5 audited that judgment; in chain B, claude-sonnet-5-5 judged and claude-opus-5-5 audited. Each model answered supported, not supported, or insufficient evidence under the same reviewer guide, with a reason code and an uncertainty flag; the auditor saw only its own chain’s judgment and accepted or revised it. No model saw the other chain, and no clinician has reviewed these answers. All 4 roles ran with adaptive thinking at medium effort, at most 8,192 output tokens for judgments and 16,384 for audits, 1 request at a time, and no caching; a rejected answer got 1 automatic retry (7 in all, plus 3 resends the investigator reviewed). One initial judgment stayed missing, so chain B covers 999 units.

**Results.** The 2 chains gave the same final decision for 960 of the 999 units both completed (96.1%): 466 of 499 in the 10 therapeutic areas and 494 of 500 in the 10 condition groups. Agreement was lowest for “Pathological conditions, signs and symptoms” (36 of 49). All 4 recorded fields matched for 802 units (80.3%). Chain A’s final decisions were 758 supported, 236 not supported, and 6 insufficient evidence; chain B’s were 784, 213, and 2. Chain A’s auditor revised 25 of 1,000 judgments (2.5%) and chain B’s 141 of 999 (14.1%); chain B’s auditor most often changed supported to not supported (30) (eTable 3).

**eFigure 1. Therapeutic-Area Classification,**

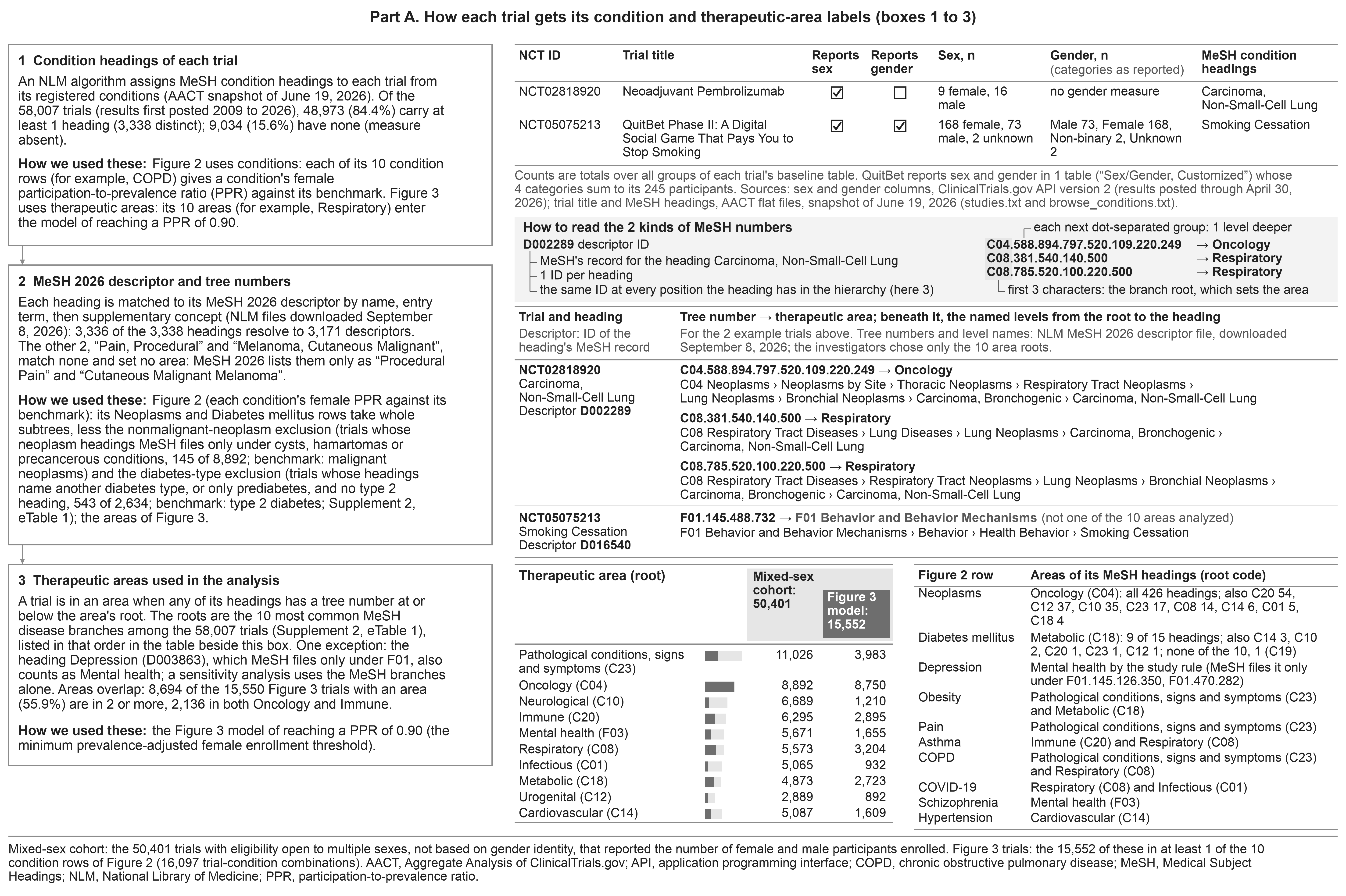

**eFigure 1 (continued).**

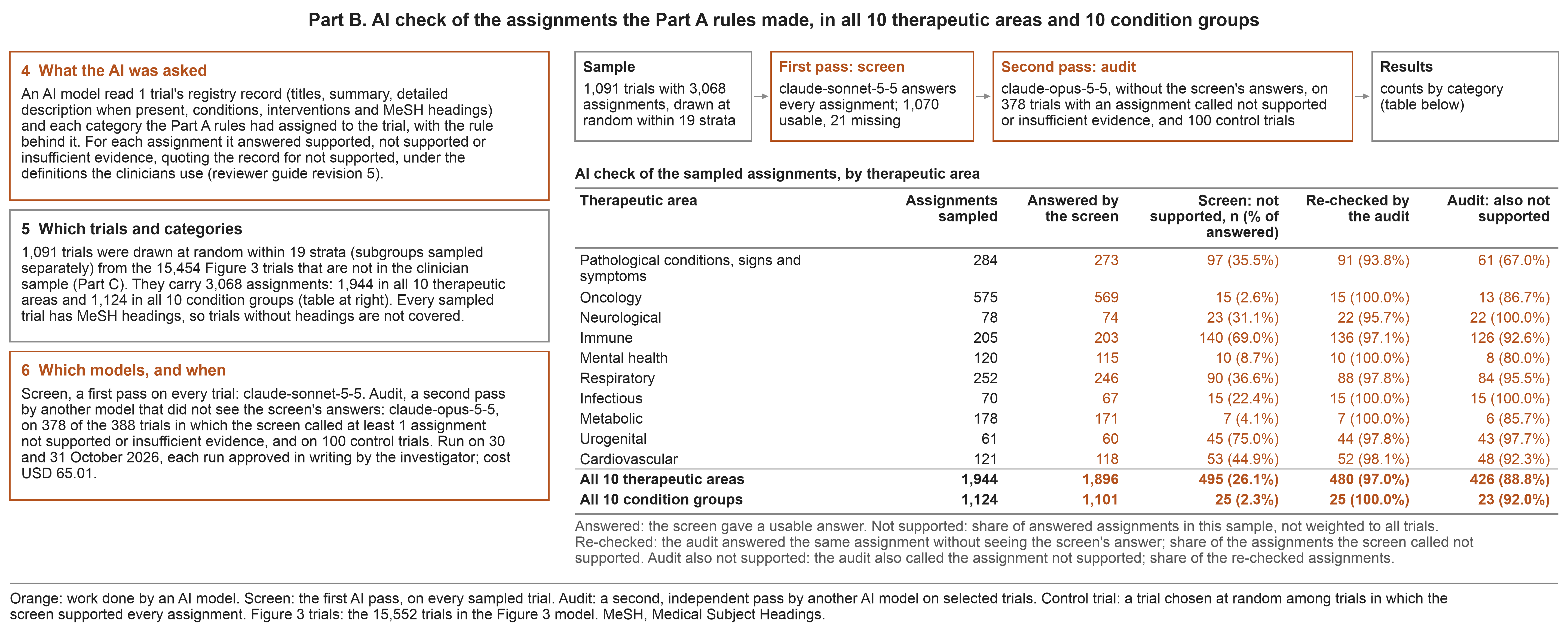

**eFigure 1 (continued).**

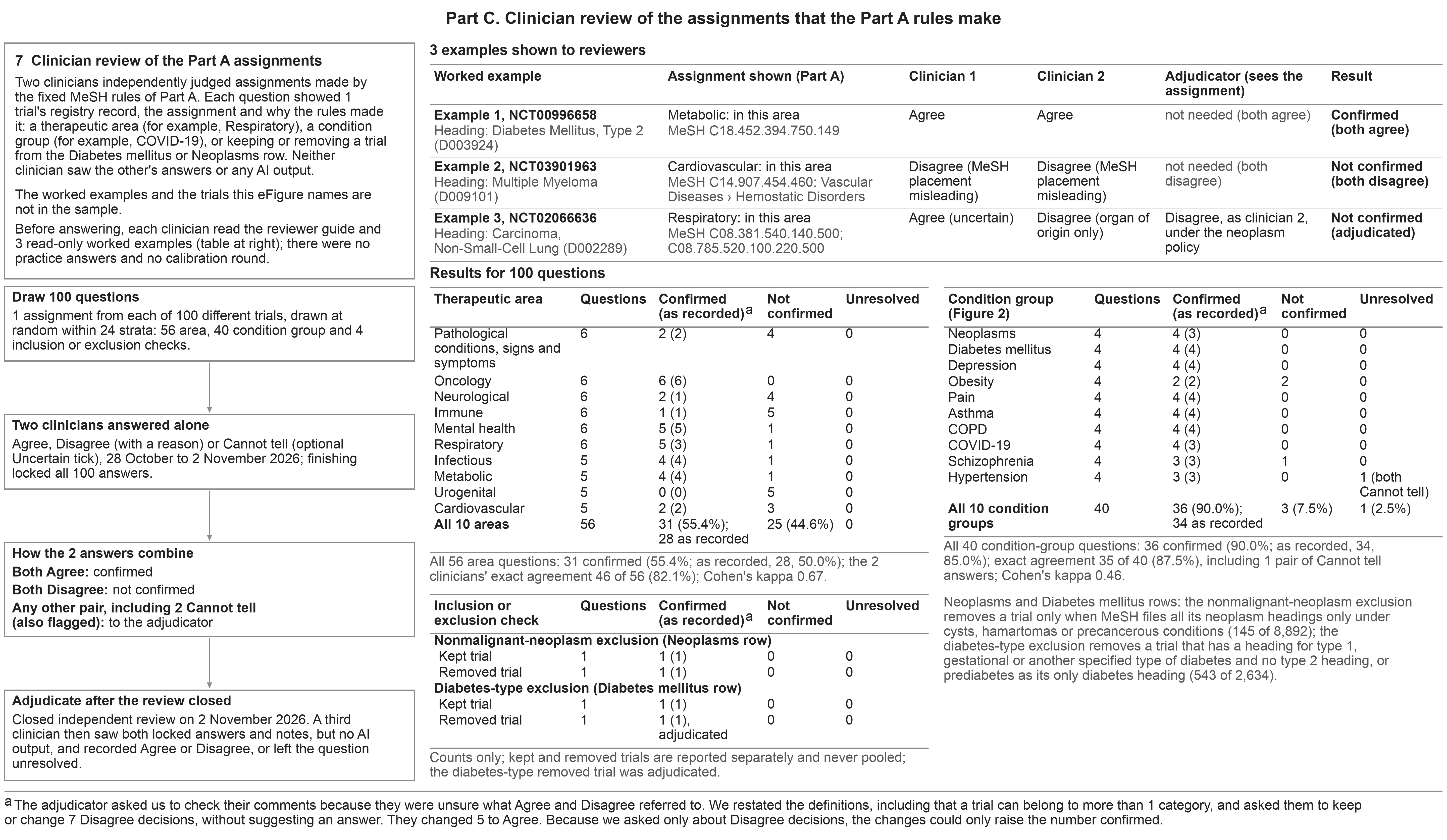

Diagrams generated with Claude Science (beta; Anthropic), running Claude Opus 5.5 and Claude Fable 5.1, using Matplotlib 3.11.1 (Python). Abbreviations: AI, artificial intelligence; COPD, chronic obstructive pulmonary disease; MeSH, Medical Subject Headings.

**eTable 1. Therapeutic Areas and Condition Categories as Parent MeSH Categories**

**A. Parent MeSH categories used as the therapeutic-area indicators**

| **Area** | **MeSH root**  **(tree number)** | **Trials, primary cohort, n (%)** | **Figure 3 model, rows (trials)** | **Distinct descriptors, n** | **Five largest contributing descriptors, trials** | **Trials entering only through descriptors also filed under another of the 10 roots,**  **n (% of area)** |
| --- | --- | --- | --- | --- | --- | --- |
| Pathological conditions, signs and symptoms | Pathological Conditions, Signs and Symptoms (C23) | 12,673 (21.8) | 4,382 (3,983) | 622 | Obesity 934; Pain 918; Pulmonary Disease, Chronic Obstructive 632; Neoplasm Metastasis 446; Postoperative Pain 441 | 6,652 (52.5) |
| Oncology | Neoplasms (C04) | 11,212 (19.3) | 8,910 (8,750) | 428 | Breast Neoplasms 1,404; Neoplasms 982; Carcinoma, Non-Small-Cell Lung 903; Prostatic Neoplasms 772; Multiple Myeloma 593 | 4,200 (37.5) |
| Neurological | Nervous System Diseases (C10) | 7,128 (12.3) | 1,291 (1,210) | 524 | Parkinson Disease 543; Alzheimer Disease 529; Stroke 511; Multiple Sclerosis 299; Migraine Disorders 269 | 4,460 (62.6) |
| Immune | Immune System Diseases (C20) | 6,618 (11.4) | 2,979 (2,895) | 179 | Asthma 655; HIV Infections 607; Multiple Myeloma 593; Diabetes Mellitus, Type 1 561; Arthritis, Rheumatoid 517 | 4,953 (74.8) |
| Mental health | Mental Disorders (F03), plus Depression (D003863), the documented exception | 6,293 (10.8) | 1,804 (1,655) | 163 | Depression 941; Schizophrenia 572; Alzheimer Disease 529; Major Depressive Disorder 519; Anxiety Disorders 477 | 1,310 (20.8) |
| Respiratory | Respiratory Tract Diseases (C08) | 5,719 (9.9) | 3,320 (3,204) | 153 | Carcinoma, Non-Small-Cell Lung 903; Asthma 655; Pulmonary Disease, Chronic Obstructive 632; COVID-19 592; Influenza, Human 511 | 4,711 (82.4) |
| Infectious | Infections (C01) | 5,660 (9.8) | 966 (932) | 330 | HIV Infections 607; COVID-19 592; Influenza, Human 511; Acquired Immunodeficiency Syndrome 458; Hepatitis C 396 | 3,006 (53.1) |
| Metabolic | Nutritional and Metabolic Diseases (C18) | 5,446 (9.4) | 3,092 (2,723) | 231 | Diabetes Mellitus, Type 2 1,459; Diabetes Mellitus 994; Obesity 934; Diabetes Mellitus, Type 1 561; Insulin Resistance 326 | 1,592 (29.2) |
| Urogenital | Urogenital Diseases (C12) | 5,414 (9.3) | 930 (892) | 250 | Prostatic Neoplasms 772; HIV Infections 607; Acquired Immunodeficiency Syndrome 458; Ovarian Neoplasms 353; Renal Insufficiency, Chronic 351 | 4,136 (76.4) |
| Cardiovascular | Cardiovascular Diseases (C14) | 5,315 (9.2) | 1,819 (1,609) | 263 | Hypertension 595; Multiple Myeloma 593; Stroke 511; Heart Failure 505; Cardiovascular Diseases 424 | 2,024 (38.1) |

A trial is in an area when any directly assigned condition heading has a MeSH 2026 tree number equal to the root or below it; a heading that resolves to the descriptor Depression (D003863; tree numbers F01.145.126.350 and F01.470.282) also places the trial in mental health. Descriptors: distinct MeSH 2026 descriptors under the root among the cohort's headings (153 to 622 per area). MeSH files a descriptor under every branch that fits it, so the areas overlap: 733 of the 2,317 descriptors that place cohort trials in an area (31.6%) are filed under 2 or more of the 10 roots, and 21,609 of the 42,622 trials in any area (50.7%) are in 2 or more areas. Abbreviations: AACT, Aggregate Analysis of ClinicalTrials.gov; MeSH, Medical Subject Headings.

**B. AI check of the Part A assignments by therapeutic area**

| **Area** | **Assignments sampled, n** | **Answered by the screen, n** | **Not supported by the screen, n (% of answered)** | **Re-checked by the audit, n** | **Also not supported by the audit, n** |
| --- | --- | --- | --- | --- | --- |
| Pathological conditions, signs and symptoms | 284 | 273 | 97 (35.5%) | 91 | 61 |
| Oncology | 575 | 569 | 15 (2.6%) | 15 | 13 |
| Neurological | 78 | 74 | 23 (31.1%) | 22 | 22 |
| Immune | 205 | 203 | 140 (69.0%) | 136 | 126 |
| Mental health | 120 | 115 | 10 (8.7%) | 10 | 8 |
| Respiratory | 252 | 246 | 90 (36.6%) | 88 | 84 |
| Infectious | 70 | 67 | 15 (22.4%) | 15 | 15 |
| Metabolic | 178 | 171 | 7 (4.1%) | 7 | 6 |
| Urogenital | 61 | 60 | 45 (75.0%) | 44 | 43 |
| Cardiovascular | 121 | 118 | 53 (44.9%) | 52 | 48 |
| **All 10 therapeutic areas** | **1,944** | **1,896** | **495 (26.1%)** | **480** | **426** |
| **All 10 condition groups** | **1,124** | **1,101** | **25 (2.3%)** | **25** | **23** |

Counts of the assignments of the 1,091 trials checked on 30 and 31 October 2026. Answered: the screen (claude-sonnet-5-5) gave a valid answer; the 21 missing screens held 48 therapeutic-area and 23 condition-group assignments. Not supported: share of answered assignments in this sample, not weighted to all trials. Re-checked: the audit (claude-opus-5-5), which did not see the screen’s answers, answered the same assignment.

**C. PPR condition categories**

| **Condition row** | **MeSH definition** | **Level** | **Rule in the main analysis** | **Mixed-sex trials, n** | **Figure 3 model, trials** |
| --- | --- | --- | --- | --- | --- |
| Neoplasms | Every heading with a tree number under Neoplasms (C04) | parent subtree | row excludes the 145 of 8,892 trials whose neoplasm headings MeSH files only under cysts, hamartomas or precancerous conditions (C04.182, C04.445 and C04.834); the benchmark is malignant cancer | 8,892 | 8,747 |
| Diabetes Mellitus | Every heading under Diabetes Mellitus (C18.452.394.750; C19.246) | parent subtree | Diabetes-type exclusion: the Diabetes Mellitus row excludes the 543 of 2,634 trials whose headings name another diabetes type, or only prediabetes, and no type 2 heading; the benchmark is type 2 diabetes | 2,634 | 2,091 |
| Depression | Heading Depression (D003863) as indexed; tree numbers F01.145.126.350, F01.470.282; MeSH files it under F01 (Behavior and Behavior Mechanisms), not Mental Disorders (F03) | single descriptor as indexed | none | 813 | 813 |
| Obesity | Heading Obesity (D009765) as indexed; tree numbers C18.654.726.750.500, C23.888.144.699.500 | single descriptor as indexed | none | 763 | 763 |
| Asthma | Heading Asthma (D001249) as indexed; tree numbers C08.127.108, C08.381.495.108, C08.674.095, C20.543.480.680.095 | single descriptor as indexed | none | 646 | 646 |
| COPD | Heading Pulmonary Disease, Chronic Obstructive (D029424) as indexed; tree numbers C08.381.495.389, C23.550.291.500.875 | single descriptor as indexed | none | 618 | 618 |
| COVID-19 | Heading COVID-19 (D000086382) as indexed; tree numbers C01.748.610.763.500, C01.925.705.500, C01.925.782.600.550.200.163, C08.381.677.807.500, C08.730.610.763.500 | single descriptor as indexed | none | 569 | 569 |
| Schizophrenia | Heading Schizophrenia (D012559) as indexed; tree numbers F03.700.750 | single descriptor as indexed | none | 559 | 559 |
| Hypertension | Heading Hypertension (D006973) as indexed; tree numbers C14.907.489 | single descriptor as indexed | benchmark compatible with caveat | 543 | 543 |
| Pain | Heading Pain (D010146) as indexed; tree numbers C23.888.592.612, F02.830.816.444, G11.561.790.444 | single descriptor as indexed | benchmark heterogenous | 748 | 748 |

Neoplasms and Diabetes Mellitus are parent subtrees narrowed only by the nonmalignant-neoplasm and diabetes-type exclusions, which exist because the benchmarks describe malignant cancer and type 2 diabetes. The other rows are single descriptors used as the registry indexes them, with no subtree expansion, so trials indexed only with a narrower descriptor are not in the row (for example, 127 mixed-sex trials with only a narrower obesity descriptor and 114 with only a narrower hypertension descriptor). The Depression heading (D003863) also places trials in mental health

Abbreviations: COPD, chronic obstructive pulmonary disease; MeSH, Medical Subject Headings; PPR, participation-to-prevalence ratio.

**D. MeSH disease branches with the most cohort trials, the basis of the 10 therapeutic areas**

| **MeSH disease branch (tree number)** | **Role in the analysis** | **Cohort trials, n (%)** | **Rank** | **Mixed-sex cohort trials, n (%)** | **Rank** | **Figure 3 model trials,**  **n (%)** | **Rank** |
| --- | --- | --- | --- | --- | --- | --- | --- |
| Pathological Conditions, Signs and Symptoms (C23) | Area: Pathological conditions, signs and symptoms | 12,673 (21.8) | 1 | 11,026 (21.9) | 1 | 3,983 (25.6) | 2 |
| Neoplasms (C04) | Area: Oncology | 11,212 (19.3) | 2 | 8,892 (17.6) | 2 | 8,750 (56.3) | 1 |
| Nervous System Diseases (C10) | Area: Neurological | 7,128 (12.3) | 3 | 6,689 (13.3) | 3 | 1,210 (7.8) | 12 |
| Immune System Diseases (C20) | Area: Immune | 6,618 (11.4) | 4 | 6,295 (12.5) | 4 | 2,895 (18.6) | 4 |
| Mental Disorders (F03) | Area: Mental health | 5,940 (10.2) | 5 | 5,376 (10.7) | 6 | 1,360 (8.7) | 10 |
| Respiratory Tract Diseases (C08) | Area: Respiratory | 5,719 (9.9) | 6 | 5,573 (11.1) | 5 | 3,204 (20.6) | 3 |
| Infections (C01) | Area: Infectious | 5,660 (9.8) | 7 | 5,065 (10.0) | 8 | 932 (6.0) | 13 |
| Nutritional and Metabolic Diseases (C18) | Area: Metabolic | 5,446 (9.4) | 8 | 4,873 (9.7) | 9 | 2,723 (17.5) | 6 |
| Urogenital Diseases (C12) | Area: Urogenital | 5,414 (9.3) | 9 | 2,889 (5.7) | 14 | 892 (5.7) | 14 |
| Cardiovascular Diseases (C14) | Area: Cardiovascular | 5,315 (9.2) | 10 | 5,087 (10.1) | 7 | 1,609 (10.3) | 9 |

Of the 58,007 cohort trials, 47,605 (82.1%) carry a heading in at least 1 ranked branch, 1,368 (2.4%) carry headings in none and 9,034 (15.6%) carry no heading.

Abbreviation: MeSH, Medical Subject Headings.

**eTable 2. Clinician Review: Independent Answers and Final Outcomes**

**A. Therapeutic areas (56 questions): the 2 clinicians’ final independent answers**

|  | **Clinician 2: Agree** | **Clinician 2: Disagree** | **Clinician 2: Cannot tell** | **Total** |
| --- | --- | --- | --- | --- |
| Clinician 1: Agree | 23 | 0 | 0 | 23 |
| Clinician 1: Disagree | 7 | 23 | 2 | 32 |
| Clinician 1: Cannot tell | 1 | 0 | 0 | 1 |
| Total | 31 | 23 | 2 | 56 |

Exact agreement 46 of 56 (82.1%); excluding the 3 pairs with a Cannot tell, 46 of 53 (86.8%); unweighted Cohen κ 0.668 (observed agreement 0.821; chance agreement 0.463).

**B. Condition groups (40 questions)**

|  | **Clinician 2: Agree** | **Clinician 2: Disagree** | **Clinician 2: Cannot tell** | **Total** |
| --- | --- | --- | --- | --- |
| Clinician 1: Agree | 33 | 0 | 1 | 34 |
| Clinician 1: Disagree | 3 | 1 | 1 | 5 |
| Clinician 1: Cannot tell | 0 | 0 | 1 | 1 |
| Total | 36 | 1 | 3 | 40 |

Exact agreement 35 of 40 (87.5%), including 1 pair of Cannot tell answers; excluding the 3 pairs with a Cannot tell, 34 of 37 (91.9%); unweighted Cohen κ 0.457 (observed agreement 0.875; chance agreement 0.770).

**C. Final outcomes**

|  | **Amended (primary)** | **As recorded (locked reference)** |
| --- | --- | --- |
| Therapeutic areas (56): Confirmed | 31 (55.4%) | 28 (50.0%) |
| Therapeutic areas (56): Not confirmed | 25 (44.6%) | 28 (50.0%) |
| Therapeutic areas (56): Unresolved | 0 (0.0%) | 0 (0.0%) |
| Therapeutic areas (56): Missing | 0 (0.0%) | 0 (0.0%) |
| Condition groups (40): Confirmed | 36 (90.0%) | 34 (85.0%) |
| Condition groups (40): Not confirmed | 3 (7.5%) | 5 (12.5%) |
| Condition groups (40): Unresolved | 1 (2.5%) | 1 (2.5%) |
| Condition groups (40): Missing | 0 (0.0%) | 0 (0.0%) |
| Inclusion or exclusion checks (4): Confirmed | 4 | 4 |
| Inclusion or exclusion checks (4): Not confirmed | 0 | 0 |
| Inclusion or exclusion checks (4): Unresolved | 0 | 0 |
| Inclusion or exclusion checks (4): Missing | 0 | 0 |

Rows are Clinician 1 and columns Clinician 2; answers are each clinician’s final independent answers (the last saved answer when the clinician finished). Cohen κ is unweighted across Agree, Disagree, and Cannot tell. As recorded: the adjudicator’s decisions as entered in the review application and locked on 3 November 2026. Amended (primary): applies the 5 changes the adjudicator confirmed after adjudication.

eTable 3. Two AI Review Chains on 1,000 Further Assignments: Final Decisions and Auditor Revisions

**A. Final decisions, chain A (rows) by chain B (columns): the 999 units both chains completed**

|  | **Chain B: supported** | **Chain B: not supported** | **Chain B: insufficient evidence** | **Total** |
| --- | --- | --- | --- | --- |
| Chain A: supported | 753 | 4 | 0 | 757 |
| Chain A: not supported | 29 | 206 | 1 | 236 |
| Chain A: insufficient evidence | 2 | 3 | 1 | 6 |
| Total | 784 | 213 | 2 | 999 |

Same final decision for 960 of 999 units (96.1%); all 4 recorded fields (decision, reason code, uncertainty flag, diabetes type) the same for 802 (80.3%).

**B. What each auditor did**

|  | **Chain A: claude-sonnet-5-5 audits claude-haiku-5-5** | **Chain B: claude-opus-5-5 audits claude-sonnet-5-5** |
| --- | --- | --- |
| Judgments audited | 1,000 | 999 |
| Accepted, n (%) | 975 (97.5) | 858 (85.9) |
| Revised, n (%) | 25 (2.5) | 141 (14.1) |
| Decision changed, n | 18 | 42 |
| Reason code changed, n | 22 | 87 |
| Uncertainty flag changed, n | 7 | 72 |

An AI auditor answered revise when at least 1 of the 4 fields changed; a revision is a change by the auditor, not a correction. Units with both chains complete: 999 in 972 trials; 1 chain B initial judgment, for a Pathological conditions, signs and symptoms unit, failed the format check twice and was closed as missing.
