## Supplement 1. Supplementary Online Content for "Sex and Gender Reporting and Female Participation Relative to Disease Burden in ClinicalTrials.gov"

eMethods 1. Cohort Construction and Data Linkage

eMethods 2. Sex and Gender Reporting States and the Extraction Audit

eMethods 3. Benchmark Derivations

eMethods 4. PPR Eligibility, Case-Definition Rules, and Models

eTable 1. Sex and Gender Reporting States

eTable 2. Cohort Derivation

eTable 3. Female Share of People With Each Condition (PPR Benchmarks)

eTable 4. Gender Identity Categories Among Trials Reporting Gender

eTable 5. Characteristics of Mixed-Sex Trials In and Outside the PPR Model

eAppendix 1. TRIPOD-LLM Reporting Checklist for the Large Language Model Steps

**eMethods 1. Cohort Construction and Data Linkage**

We linked studies with posted results from the ClinicalTrials.gov API (version 2) to the AACT flat-file export on the NCT identifier. For each completion month, we took the share of cohort trials completed in 2016 through 2019 whose results posted within the time that month had before April 30, 2026, and averaged over months, weighted as in the reference years. The estimates were 11.0% for 2025 and 0.3% for 2026 (15.3% and 0.5% with 2019 through 2020 as the reference).

**eMethods 2. Sex and Gender Reporting States and the Extraction Audit**

Three states are kept apart and never combined: (1) a breakdown is reported; (2) participants are recorded only as unknown or not reported, or the registrant states that sex was not collected; (3) the measure is absent. Aggregate or all zero measures are counted separately. Participants recorded as unknown inside a reported breakdown stay as unknown (532 trials). The mixed-sex cohort is the 50,448 trials open to multiple sexes with a female and male breakdown, less 47 trials whose breakdown counts units other than participants, such as eyes, lesions or samples.

**Reporting rules.**

Reporting sex and providing a complete, usable female and male distribution are kept apart. Female and male counts that cover only some participants (partial arm-level or cluster-level counts) are not used as a distribution.

**Extraction audit.**

Our extraction pipeline flagged 575 trials whose baseline sex or gender table had a customized or non-standard structure, where totals can double count participants across arms (all had two or more categories with denominators; 178 were titled “Sex/Gender, Customized”; 519 are in the primary cohort). A large language model made a first pass at extracting information from each trial. The model was Claude Haiku 4.5 (Anthropic), accessed May 2026 by M.A. One reviewer (B.Y., supervised by M.G.) checked every parse against the registry record, rating format and numbers separately, so no parse entered the analysis unchecked. Of the 575 parses, 475 (82.6%) were correct and 100 had a format or count problem. The reviewer supplied corrected values for 98 of the 100 and confirmed them, and the analyses apply them where the extract is read. The other 2 had written notes only (one parse with a format problem but correct counts, and one registry record that is internally inconsistent) and kept their extracted values. For one corrected trial that reports patients and providers in the same table, the analyses count patients only. One trial’s gender categories sum to more than 110% of its enrollment and was excluded (eTable 5).

**eMethods 3. Benchmark Derivations**

The PPR denominator is the female share of people with the condition, not prevalence among women: from sex-specific crude prevalence, (prevalence in women × women) / (prevalence in women × women + prevalence in men × men).

For 8 conditions the benchmark is the female share of diagnosed patients in PCORnet. Deviations are below.

**COPD:** adults in the 2023 National Health Interview Survey ever told they had COPD, emphysema or chronic bronchitis (crude prevalence 4.78% in women, 3.81% in men) give weighted case counts of 6,312,323 women and 4,799,338 men, a female share of 56.8%.

**Hypertension:** the 2023 PCORnet query cohort with diagnosed hypertension gives 4,490,038 / (4,490,038 + 4,136,524) = 52.0%.

**HIV:** for treatment trials, the benchmark is the female share of persons aged 13 years or older living with diagnosed HIV at year-end 2023 in the National HIV Surveillance System, 252,004 / (252,004 + 862,584) = **22.6%**.

**Pain:** a single heterogeneous heading, against the chronic-pain benchmark.

**eMethods 4. PPR Eligibility, Case-Definition Rules, and Models**

**Condition categories.** Conditions come from each trial's MeSH headings, resolved to MeSH 2026 descriptors. Headings in the Neoplasms subtree (C04) form one category, headings in the Diabetes Mellitus subtrees (C18.452.394.750 and C19.246) another. Every other heading forms its own category. Figure 2 and the regression count a trial in every category it carries; Table 1 presents each trial in its single most prevalent category.

**Eligibility rule.** A single rule defines the PPR analysis for Figure 2 and the regression: the 10 categories with the most mixed-sex trials (counting a trial in every category it carries) enter, each against its benchmark (eTable 3). The top 10 categories give us 16,097 trial-condition rows from 15,552 trials (487 trials contribute more than one row).

**Diabetes-type exclusion.** Against a type 2 diabetes benchmark, the diabetes row drops trials whose MeSH 2026 headings name another diabetes type or only prediabetes and no type 2 heading: 543 of 2,634 (496 type 1 only, 44 prediabetes only, 2 latent autoimmune diabetes in adults and 1 gestational only).

**Nonmalignant-neoplasm exclusion.** The neoplasms row drops trials whose Neoplasms headings are all on a deterministic list of cysts, precancerous conditions and hamartomas (145 of 8,892).

The model outcome is whether a trial-condition row reaches a PPR of at least 0.90. Logistic regression adjusts for funder type, phase, US sites, primary completion year (centered at 2009), log enrollment, number of countries and the area indicators, with errors clustered by trial; medians carry bootstrap percentile 95% CIs (2,000 replicates). The therapeutic-area indicators are the 10 most common MeSH disease branches among the 58,007 cohort trials. One documented exception: the descriptor Depression (D003863), which MeSH files under Behavior and Behavior Mechanisms (F01) rather than Mental Disorders (F03), also places a trial in mental health.

**eTable 1. Sex and Gender Reporting States**

**A. Reporting states as classified by the pipeline, before hand verification**

| **State** | **Trials, n (%)** |
| --- | --- |
| Sex only | 57,157 (98.53%) |
| Both sex and gender | 515 (0.89%) |
| Gender only | 38 (0.07%) |
| Any breakdown reported | 57,710 (99.49%) |
| Participants recorded only as unknown or not reported, or a statement that sex was not collected | 35 (0.06%) |
| Measure absent | 3 (0.01%) |
| Measure present without a usable breakdown (aggregate or all-zero counts) | 259 (0.45%) |

Primary cohort (58,007 trials).

**B. Final reporting states by funder type**

| **State or quantity** | **Industry** | **NIH** | **Government (US federal and other public)** | **Other** |
| --- | --- | --- | --- | --- |
| Trials, n (column total) | 30,922 | 9,650 | 2,532 | 14,903 |
| Sex only | 30,787 (99.6) | 9,320 (97) | 2,465 (97) | 14,596 (98) |
| Both sex and gender | 46 (0.15) | 277 (2.9) | 32 (1.3) | 160 (1.1) |
| Gender only | 2 (0.01) | 20 (0.21) | 4 (0.16) | 12 (0.08) |
| Neither sex nor gender | 87 (0.28) | 33 (0.34) | 31 (1.2) | 135 (0.91) |
| Participants recorded only as unknown or not reported, or sex stated as not collected | 44 (0.14) | 19 (0.20) | 22 (0.87) | 88 (0.59) |
| No sex or gender measure | 24 (0.08) | 10 (0.10) | 6 (0.24) | 31 (0.21) |
| Measure present without a usable breakdown | 19 (0.06) | 4 (0.04) | 3 (0.12) | 16 (0.11) |
| Reported sex (sex only plus both) | 30,833 (99.7) | 9,597 (99.5) | 2,497 (99) | 14,756 (99.0) |
| Reported gender (gender only plus both) | 48 (0.16) | 297 (3.1) | 36 (1.4) | 172 (1.2) |
| Incomplete-composition flag (female and male counts cover only some participants) | 7 (0.02) | 3 (0.03) | 2 (0.08) | 2 (0.01) |
| Values and an explicit unknown in the same measure | 102 (0.33) | 264 (2.7) | 37 (1.5) | 129 (0.87) |

Percentages of the column total.

Abbreviations: FDAAA, Food and Drug Administration Amendments Act; NIH, National Institutes of Health.

**eTable 2. Cohort Derivation**

| **Step** | **n** |
| --- | --- |
| All AACT study records | 590,350 |
| Interventional | 450,483 |
| With posted results | 74,074 |
| Primary completion on or after September 27, 2008 | 69,788 |
| Not terminated | 58,763 |
| Excluded: results first posted before January 1, 2009, or after April 30, 2026 | 756 |
| Primary cohort (all analyses): interventional trials with posted results, primary completion on or after September 27, 2008, not terminated, results first posted January 1, 2009, through April 30, 2026 | 58,007 |

Exclusions are the differences between consecutive rows, except the exclusion row: 756 trials with results first posted outside the study window (1 before January 1, 2009, and 755 after April 30, 2026).

Abbreviations: AACT, Aggregate Analysis of ClinicalTrials.gov; FDAAA, Food and Drug Administration Amendments Act.

**eTable 3. Female Share of People With Each Condition (PPR Benchmarks)**

| **Condition row** | **Female share, %** | **Source; period** | **Case definition** |
| --- | --- | --- | --- |
| Neoplasms | 52.1 | PCORnet Population Insights report, malignant cancer; 2025 encounter year | At least 1 face-to-face encounter at a PCORnet site in 2025 and a diagnosis code for any malignant cancer on a PCORnet list within 2021-2025 |
| Diabetes Mellitus | 51.9 | PCORnet Population Insights report, metabolic and endocrine conditions; 2024 encounter year | At least 1 face-to-face ambulatory encounter at a PCORnet site in 2024 and a diagnosis code for the condition within 2020-2024 |
| Depression | 69.3 | PCORnet network-wide query; 2023 | PCORnet code list 'Diagnosis Codes for Depression' |
| Obesity | 61.1 | PCORnet Population Insights report, metabolic and endocrine conditions; 2024 encounter year | At least 1 face-to-face ambulatory encounter at a PCORnet site in 2024 and a diagnosis code for the condition within 2020-2024 |
| Asthma | 60.2 | PCORnet network-wide query; 2023 | PCORnet code list 'Diagnosis Codes for Asthma': 26 ICD-10-CM J45.x codes and 20 SNOMED CT concepts |
| Pulmonary Disease, Chronic Obstructive (COPD) | 56.8 | National Health Interview Survey, Sample Adult file; 2023 | Yes to: 'Have you EVER been told by a doctor or other health professional that you had chronic obstructive pulmonary disease, COPD, emphysema, or... |
| COVID-19 | 60.1 | PCORnet network-wide query; 2023 | PCORnet code list 'Diagnosis Codes for Covid-19': ICD-10-CM U07.1 and B97.29, SNOMED CT 840539006 and 27619001 |
| Schizophrenia | 42.0 | PCORnet data report, mental and behavioral health conditions; 2021-01-01 to 2022-12-31 | At least 2 encounters ≥60 days apart and ≥1 schizophrenia diagnosis code at a PCORnet site during 2021-2022 |
| Hypertension | 52.0 | PCORnet network-wide query; 2023 | PCORnet code list 'Diagnosis Codes for Hypertension' |
| Pain | 53.3 | National Health Interview Survey, Sample Adult file; 2023 | 'In the past three months, how often did you have pain?' answered 'most days' or 'every day'; the heading also includes acute, postoperative and procedural pain trials |
| Chronic pain subset (Chronic Pain or Intractable Pain headings) | 53.3 | National Health Interview Survey, Sample Adult file; 2023 | 'In the past three months, how often did you have pain?' answered 'most days' or 'every day' |
| HIV Infections, treatment trials | 22.6 | National HIV Surveillance System; Year-end 2023 | Persons living with diagnosed HIV infection |

Female share: the female proportion of people with the condition in the source population (the PPR denominator). Case definitions are abridged from each source’s own documentation: the PCORnet Population Insights and data reports and the code lists of the PCORnet network-wide queries, the 2023 NHIS Sample Adult questionnaire items, and the National HIV Surveillance System definition of diagnosed HIV. Each female share is the source’s female count divided by its female plus male counts (survey-weighted for NHIS). COPD, chronic obstructive pulmonary disease; NHIS, National Health Interview Survey; PPR, participation-to-prevalence ratio.

**eTable 4. Gender Identity Categories Among Trials Reporting Gender**

| **Category** | **Participants, n** | **Trials with a nonzero count, n** | **Trials posting the category, n (%)** |
| --- | --- | --- | --- |
| Transgender, with or without a sex term | 4,473 | 199 | 203 (37.7) |
| of which the label also names a sex  (for example 'transgender woman') | 3,058 | 107 | 108 (20.1) |
| of which no sex term | 1,415 | 97 | 100 (18.6) |
| Non-binary or gender-expansive | 4,711 | 222 | 223 (41.4) |
| Questioning | N<25 | n<25 | n<25 |
| Combined transgender or gender-diverse group | 291 | n<25 | n<25 |
| Another or self-described gender (includes bare 'Other' rows) | 2,613 | 215 | 222 (41.3) |
| Total, gender identity categories | 12,105 | 524 | 525 (97.6) |
| Intersex | N<25 | N<25 | 13 (2.4) |
| Cisgender-labeled | 19,114 | 38 | 38 (6.9) |
| Explicit unknown, not reported or declined | 11,673 | 154 | 155 (28.0) |

Participants are summed over trial by category cells (each category counted once per trial, at its largest value across the trial's count-driving baseline tables). The gender identity categories exclude 15 trials whose counts were not participant counts (3) or whose category sum exceeded enrollment by more than 10% (12); percentages use the remaining 538 trials, and the separately reported rows use all 553. 'Another or self-described gender' includes rows labeled only 'Other'. Explicit unknown=not reported or declined is a recorded value and is distinct from a trial with no gender measure.

**eTable 5. Characteristics of Mixed-Sex Trials In and Outside the PPR Model**

| **Characteristic** | **A. In the PPR model**  **(n = 15,552)** | **B. Top 10 condition, not in the model**  **(n = 651)** | **C. No top 10 condition**  **(n = 34,198)** | **SMD, A vs B and C** |
| --- | --- | --- | --- | --- |
| Trial-condition rows in the PPR model; trials with more than one row | 16,097; 487 | not applicable | not applicable |  |
| Funder type: NIH | 3,417 (22.0) | 120 (18.4) | 4,577 (13.4) | 0.22 |
| Trial phase: N/A or other | 3,435 (22.1) | 272 (41.8) | 12,625 (36.9) | -0.33 |
| Trial phase: Phase 1/2 | 1,240 (8.0) | 24 (3.7) | 1,264 (3.7) | 0.18 |
| Trial phase: Phase 2 | 5,385 (34.6) | 119 (18.3) | 6,585 (19.3) | 0.35 |
| Trial phase: Phase 4 | 1,229 (7.9) | 53 (8.1) | 3,991 (11.7) | -0.13 |
| US study sites | 12,234 (78.7) | 510 (78.3) | 24,413 (71.4) | 0.17 |
| Child-eligible | 1,872 (12.0) | 212 (32.6) | 6,792 (19.9) | -0.22 |
| Registry primary purpose: other | 1,590 (10.2) | 116 (17.8) | 5,657 (16.5) | -0.19 |
| Registry primary purpose: prevention | 773 (5.0) | 35 (5.4) | 3,702 (10.8) | -0.22 |
| Registry primary purpose: treatment | 12,478 (80.2) | 469 (72.0) | 23,324 (68.2) | 0.28 |
| No. of countries, median (IQR) | 1 (1 to 1) | 1 (1 to 1) | 1 (1 to 1) | 0.12 |
| Percent female, median (IQR) | 45.0 (33.3 to 57.1) | 46.5 (36.1 to 56.4) | 49.5 (35.2 to 62.5) | -0.13 |

Rows shown are the 12 characteristic levels whose absolute standardized mean difference (SMD) is at least 0.10, the conventional imbalance threshold. Values are n (%) or median (IQR).

Abbreviations: IQR, interquartile range; N/A, not applicable; NIH, National Institutes of Health; PPR, participation-to-prevalence ratio

**eAppendix 1. TRIPOD-LLM Reporting Checklist for the Large Language Model Steps**

The study used large language models (LLMs) in 2 steps. Step 1, extraction support: Claude Haiku 4.5 made a first pass at 575 complex baseline sex or gender tables, and a reviewer checked every one (eMethods 2); only this step feeds the analyses, through the checked values. Step 2, checks of the MeSH-rule assignments: on 30 and 31 October 2026, claude-sonnet-5-5 checked 3,068 assignments in 1,091 trials and claude-opus-5-5 re-checked selected trials; on 3 and 4 November 2026, 2 review chains (claude-haiku-5-5 audited by claude-sonnet-5-5; claude-sonnet-5-5 audited by claude-opus-5-5) judged 1,000 further assignments. No estimate uses the step 2 results (Supplement 2, eMethods, section 2; eFigure 1, Part B; eTable 3).

| **Item** | **TRIPOD-LLM item (abridged)** | **Step 1: extraction support** | **Step 2: checks of the assignments** |
| --- | --- | --- | --- |
| 1 | Title identifies the study as developing or evaluating an LLM, with task, population and outcome | Not reported: the LLM was a support tool, not the study subject | Not reported: the LLMs were support tools, not the study subject |
| 2 | Abstract (TRIPOD-LLM for abstracts) | Not reported: the abstract does not describe this step | Not reported: the abstract does not describe this step |
| 3a | Healthcare context and rationale for using the LLM | Methods, Trial dataset and data extraction; eMethods 2 | Supplement 2, eMethods, opening paragraph and section 2 |
| 3b | Target population, intended use and intended users | Methods; eMethods 2 | Supplement 2, eMethods, section 2 |
| 4 | Study objectives, including the stage (development, tuning or validation) | eMethods 2 | Supplement 2, eMethods, section 2 |
| 5a | Sources of data and the rationale for using them | eMethods 2 | Supplement 2, eMethods, section 2 |
| 5b | Description of the data and their distribution | eMethods 2 | Supplement 2, eMethods, section 2; eTable 1, panel B |
| 5c | Dates of the oldest and newest text | Methods: results posted 2009 to 2026 | Partly: cohort dates only (Methods) |
| 5d | Data preprocessing and quality checks | Not reported: how the tables were presented to the model | Supplement 2, eMethods, section 2 |
| 5e | Handling of missing and imbalanced data | eMethods 2 | Supplement 2, eMethods, section 2 |
| 6a | LLM name, version and last training date | Methods (Claude Haiku 4.5, accessed May 2026); training date not published by the developer | Supplement 2, eMethods, section 2 (claude-sonnet-5-5, claude-opus-5-5, claude-haiku-5-5) |
| 6c | How text was generated: prompt engineering and inference settings | Not reported | Supplement 2, eMethods, section 2; settings of the October check and the prompts not reported |
| 6d | Initial and postprocessed output | eMethods 2 | Supplement 2, eMethods, section 2 |
| 6e | Classification rules and thresholds | Not applicable: extraction, not classification | Supplement 2, eMethods, section 2 ( |
| 7b | Relevance of the outcome metrics to the task | eMethods 2 | Supplement 2, eTable 1; eMethods, section 2 |
| 7c | Outcome definition, how predictions were calculated, inference dates and metrics | Methods; eMethods 2 ( | Supplement 2, eMethods, section 2 (30 and 31 October 2026; 3 and 4 November 2026; counts and agreement) |
| 7d | Qualifications, instructions and agreement of human assessors | Partly: eMethods 2 (1 reviewer, B.Y.); training and instructions not stated | Not applicable: no human assessors |
| 7e | Comparison with other LLMs, humans or benchmarks | eMethods 2 (a reviewer checked every parse) | Supplement 2, eMethods, section 2 |
| 8a | Labeling guidelines with examples | Partly: eMethods 2 (review rules summarized; no examples) | Supplement 2, eMethods, section 2 |
| 8b | Number of labelers and interannotator agreement | eMethods 2 | Supplement 2, eMethods, section 2 |
| 8c | Background of labelers or characteristics of labeling models | Not reported: reviewer background | Supplement 2, eMethods, section 2 (model identifiers) |
| 9a | Prompt design, curation and selection | Not reported | Partly: Supplement 2, eMethods, section 2 |
| 9b | Data used to develop the prompts | Not reported | Not reported |
| 12 | Compute or cost | Not reported | Supplement 2, eFigure 1, Part B (USD 65.01), for the check of 30 and 31 October 2026; the 2 review chains of 3 and 4 November 2026 cost USD 133.49 |
| 13 | Ethics approval | Methods (Duke IRB exemption) | Methods |
| 14a | Funding and role of the funder | Funding/Support; Role of the Funder/Sponsor | Funding/Support; Role of the Funder/Sponsor |
| 14b | Conflicts of interest | Conflict of Interest Disclosures | Conflict of Interest Disclosures |
| 14e | Availability of the study data | Data Sharing Statement | Data Sharing Statement |
| 14f | Availability of the code | Not stated for this step | Not stated |
| 17 | Performance by prespecified metrics or human evaluation | Methods; eMethods 2 (475 of 575 correct) | Supplement 2, eMethods, section 2; eTable 1, panel B; eTable 3 |
| 19a | Interpretation of the main results | eMethods 2 | Supplement 2, eMethods, section 2 |
| 19b | Limitations | Not reported in the Limitations | Partly: Supplement 2, eTable 1, panel B note (consistency, not accuracy; not weighted to all trials); eMethods, section 2 |
| 19c | Known data challenges for the task | eMethods 2 | Supplement 2, eMethods, section 2 (models saw the displayed assignment and the rule behind it) |
| 19d | Intended use, end user and level of human oversight | eMethods 2 | Supplement 2, eMethods, section 2 |
| 19e | Handling of poor or missing input | Partly: eMethods 2 | Supplement 2, eMethods, section 2 |
| 19f | Expertise required of users | Not stated | Not stated |
| 19g | Next steps | Not reported | Not reported |
| 2a to 2l | Abstract items | Not reported: the abstract does not describe this step | Not reported: the abstract does not describe this step |
| 6b, 7a, 10, 11, 14c, 14d, 15, 16a to 16d, 18 | Items for LLM development or tuning, generative outputs, summarization, protocols and registration, public involvement, patient or EHR data flow, and LLM updating | Not applicable: existing models used as released; no tuning, summarization, patient-level data or updates; no protocol or registration for this step; no patient or public involvement | Not applicable: as for step 1 |
